# Resting-state functional connectivity in children cooled for neonatal encephalopathy

**DOI:** 10.1101/2022.12.17.22283576

**Authors:** Arthur P C Spencer, Marc Goodfellow, Ela Chakkarapani, Jonathan C W Brooks

**Author notes:** These authors contributed equally to this work (Ela Chakkarapani, Jonathan C W Brooks). Corresponding Author: Arthur P C Spencer; Centre de Recherche en Radiologie PET3, CHUV, Rue du Bugnon 46, 1011 Lausanne, Switzerland.

## Abstract

Therapeutic hypothermia improves outcomes following neonatal hypoxic-ischaemic encephalopathy (HIE), reducing cases of death and severe disability such as cerebral palsy compared to normothermia management. However, when cooled children reach early school-age they have cognitive and motor impairments which are associated with underlying alterations to brain structure and white matter connectivity. It is unknown whether these differences in structural connectivity are associated with differences in functional connectivity between cooled children and healthy controls. Resting-state fMRI has been used to characterise static and dynamic functional connectivity in children, both with typical development and those with neurodevelopmental disorders. Previous studies of resting-state brain networks in children with HIE have focussed on the neonatal period. In this study, we used resting-state fMRI to investigate static and dynamic functional connectivity in children aged 6-8 years who were cooled for neonatal HIE without cerebral palsy (n = 22, median age [IQR] 7.08 [6.85-7.52] years), and healthy controls matched for age, sex and socioeconomic status (n = 20, median age [IQR] 6.75 [6.48-7.25] years). Using group independent component analysis, we identified 31 intrinsic functional connectivity networks consistent with those previously reported in children and adults. We found no case-control differences in the spatial maps of these intrinsic connectivity networks. We constructed subject-specific static functional connectivity networks by measuring pairwise Pearson correlations between component time courses, and found no case-control differences in functional connectivity after FDR correction. To study the time-varying organisation of resting-state networks, we used sliding-window correlations and deep clustering to investigate dynamic functional connectivity characteristics. We found k = 4 repetitively occurring functional connectivity states, which exhibited no case-control differences in dwell time, fractional occupancy, or state functional connectivity matrices. In this small cohort, the spatiotemporal characteristics of resting-state brain networks in cooled children without severe disability were too subtle to be differentiated from healthy controls at early school-age, despite underlying differences in brain structure and white matter connectivity, possibly reflecting a level of recovery of healthy resting-state brain function. To our knowledge, this is the first study to investigate resting-state functional connectivity in children with HIE beyond the neonatal period, and the first to investigate dynamic functional connectivity in any children with HIE.

## Introduction

Therapeutic hypothermia has considerably improved outcomes following neonatal hypoxic-ischaemic encephalopathy (HIE) secondary to perinatal asphyxia. Cooled infants are at reduced risk of death or severe disability, such as cerebral palsy, compared to normothermia management following HIE (Jacobs et al., 2013; Shankaran et al., 2012; Thoresen et al., 2021). Therapeutic hypothermia is therefore standard care for HIE in most high-income counties. However, despite the benefits of therapeutic hypothermia, there are still aspects of brain development which are impacted by HIE. At early school-age, children cooled for HIE, who do not have cerebral palsy, have cognitive and motor impairments (Jary et al., 2019; Lee-Kelland et al., 2020), attention and visuospatial processing difficulties (Tonks et al., 2019), and communication difficulties (Robb et al., 2022) compared to healthy controls. An understanding of the differences in brain structure and function between cooled children and healthy controls is required to inform research into therapeutic intervention strategies to promote healthy brain development.

Functional magnetic resonance imaging (fMRI) allows non-invasive investigation of brain activity by measuring changes in the blood oxygen level dependent (BOLD) signal. In resting-state fMRI, the participant is scanned during rest (i.e. without engaging in a task or responding to stimuli) in order to measure spontaneous fluctuations in the BOLD signal (Biswal et al., 1995). Functional connectivity (FC) analysis allows investigation of functional interactions across the brain, by measuring correlations between pairs of brain regions in these low-frequency fluctuations of recorded BOLD signal (Lee et al., 2013; Smith et al., 2013; van den Heuvel and Pol, 2010). This approach can be extended to study the time-varying organisation of resting-state brain activity using dynamic functional connectivity (dFC) analysis (Calhoun et al., 2014; Cohen, 2018; Hutchison et al., 2013; Preti et al., 2017). One such approach is to use sliding-window correlations to calculate a series of FC matrices for each subject, which can then be clustered at the group level, revealing brain states representing repetitively occurring FC patterns (Allen et al., 2014; Calhoun et al., 2014). Spatiotemporal characteristics of these dFC states have been characterised in typically developing children (Marusak et al., 2017; Rashid et al., 2018), and have been shown to be sensitive to neurodevelopmental outcomes (Harlalka et al., 2019; He et al., 2018; Li et al., 2020; Rashid et al., 2018, 2014; Zhu et al., 2023).

Studies of neonates with HIE (including both those with and without severe disability such as cerebral palsy) have found alterations to resting-state FC compared to healthy controls (Jiang et al., 2022; Tusor, 2014), and associations between FC and HIE severity (Boerwinkle et al., 2022; Li et al., 2019). However, it is unclear whether these alterations in the neonatal period affect brain function in later life. We have previously shown that children cooled for HIE have disrupted white matter connectivity (Byrne et al., 2023; Spencer et al., 2021b, 2021a) and structural alterations to subcortical structures (Spencer et al., 2023) and mammillary bodies (Spencer et al., 2022) compared to healthy controls at early school-age. It is unknown whether these alterations to brain structure and structural connectivity are associated with measurable differences in functional brain activity in these children.

In this study, we investigated resting-state brain activity using fMRI in children aged 6-8 years without cerebral palsy who were treated with therapeutic hypothermia for neonatal HIE (cases), and healthy controls matched for age, sex and socioeconomic status. We used group-level independent component analysis (ICA) to determine a set of intrinsic connectivity networks (ICNs), then studied case-control differences in the spatial maps of these ICNs, and in static and dynamic FC between ICNs. To our knowledge, this is the first study to investigate FC in children with HIE beyond the neonatal period, and the first study to investigate dFC in any children with HIE.

## Methods

### Participants

This study investigated participants of the ‘CoolMRI’ study (Lee-Kelland et al., 2020; Spencer et al., 2021b), a study of early school-age children without cerebral palsy who received therapeutic hypothermia as a neuroprotective intervention for neonatal HIE, and control children matched for age, sex, and socioeconomic status. Informed and written consent was obtained from the parents of participants and assent obtained from the children. Ethical approval was obtained from the North Bristol Research Ethics Committee and the Health Research Authority (REC ID: 15/SW/0148).

Cases were aged 6-8 years and were sequentially selected from those who received therapeutic hypothermia between October 2007 and November 2012 for moderate to severe encephalopathy, confirmed by amplitude-integrated EEG assessment (Thoresen et al., 2021), secondary to perinatal asphyxia. Cases did not have a diagnosis of cerebral palsy at 2 and at 6-8 years based on neurological examination and assessment of motor function. Children were excluded if they were cooled outside the standard criteria, born before 35 weeks gestation, had any additional diagnosis apart from HIE (such as genetic or metabolic disorder), had a major intracranial haemorrhage or congenital brain malformation visible on neonatal MRI, or were non-native English speakers.

Age, sex and socioeconomic status matched controls were recruited through local schools and newsletters circulated at the University of Bristol. Children were included who were born at >35 weeks gestation, had not had perinatal asphyxia with HIE and spoke English as their primary spoken language.

Socioeconomic status was measured based on participant’s postcode at examination, using the index of multiple deprivation as defined for England by the UK Government (www.gov.uk/government/statistics/english-indices-of-deprivation-2019). Each postcode in England is assigned a number, on a scale of 1–10, indicating the decile within which the local area is ranked in the country, from most deprived (1) to least deprived (10).

### MRI acquisition

Images were acquired using a 3 tesla Siemens Magnetom Skyra and a 32-channel receive-only head coil. A child-friendly, detailed explanatory video was sent to the family before assessment day and presented again on the day of the scan together with the typical sounds in the MRI scanner. Head movement was minimised using cushions. A T1-weighted volumetric scan was obtained, for spatial normalisation, with a magnetisation-prepared rapid acquisition gradient echo (MPRAGE) pulse sequence using the following parameters: echo time (TE) = 2.19 ms; inversion time (TI) = 800 ms; repetition time (TR) = 1500 ms; flip angle = 9°; field of view = 234 x 250 mm; 176 slices; 1.0 mm isotropic voxels; GeneRalized Autocalibrating Partially Parallel Acquisitions (GRAPPA) acceleration factor 4 (Griswold et al., 2002). During acquisition of the volumetric scan, a film of the participants’ choice was projected onto a screen visible through the mirror assembly of the head coil. During the resting-state functional acquisition, the film was turned off and participants were instructed to keep their eyes open and look at a central fixation cross. T2*-weighted functional images were acquired using a gradient echo planar imaging sequence with the following parameters: TE = 30 ms; TR = 906 ms; multiband factor 6; flip angle = 60°; field of view = 185 x 185 mm; matrix = 64 x 64; slice thickness = 3.125 mm; 36 slices; 2.890 x 2.890 x 3.125 mm voxels. We acquired 300 volumes giving a scan time of 4 minutes 32 seconds. We also acquired dual-(gradient)-echo images for distortion correction of fMRI data (see below).

### Preprocessing

Resting-state fMRI data were preprocessed using FEAT (Woolrich et al., 2001) from the FMRIB Software Library (FSL v6.0, https://fsl.fmrib.ox.ac.uk) (Jenkinson et al., 2012; Smith et al., 2004). Processing steps were as follows: a) the first 5 volumes in the sequence were discarded to ensure steady-state magnetisation, leaving 295 volumes (4 minutes 27 seconds); b) motion correction was then applied with MCFLIRT (Jenkinson et al., 2002) to align all volumes in the sequence using rigid-body registration; c) the derived fieldmap was used to correct distortions (induced by magnetic field inhomogeneities) in the fMRI data; d) non-brain tissue was removed using BET; e) Spatial smoothing was performed with a 5 mm full-width at half-maximum Gaussian kernel; f) highpass temporal filtering was applied with a cutoff of 150 s to remove low-frequency artefacts. Preprocessed fMRI data were then transformed to Montreal Neurological Institute (MNI) standard space; first, subject fMRI data were registered to the subject’s T1-weighted image using rigid-body registration, then the subject T1-weighted image was registered to the MNI standard template using nonlinear registration and the resulting transformation was applied to the fMRI data.

Following standard preprocessing steps, each subject’s fMRI data was cleaned to remove artefacts due to motion, physiological noise, and scanner noise, using FSL’s FIX (Griffanti et al., 2014; Salimi-Khorshidi et al., 2014). FIX uses a training dataset to automatically classify subject-level ICA components (calculated using MELODIC from FSL) into signal and noise, then regresses out the noise components from the fMRI data. A study-specific training dataset was generated by hand-labelling components from a random sample of 15 subjects which were matched to the full cohort for case-control status. For each subject in the training sample, components were labelled signal or noise by two raters (APCS & JCWB) based on characteristics of the spatial maps, timeseries, and frequency spectra (for detailed description of characteristics of signal and noise components, see (Griffanti et al., 2017)). Leave-one-out cross validation of the training dataset gave a mean true positive rate of 94.2% and a mean true negative rate of 88.4%. The training dataset was used to denoise all subjects’ fMRI data, including regressing out the movement parameters estimated during the motion correction preprocessing step.

### Quality Control

To assess quality of the fMRI scan, we quantified the amount of movement of each subject during acquisition using mean framewise displacement and maximum absolute displacement. Framewise displacement combines measurements of translation (x, y, z) and rotation (pitch, yaw, roll) into a single scalar quantity to summarise instantaneous head motion at each time point. This was calculated according to (Power et al., 2012), using the movement parameters estimated during the motion correction preprocessing step, and averaged across time points to give mean framewise displacement for each subject. Note that this is likely an overestimation of the framewise displacement, as rotational displacements are calculated based on an approximate radius (distance from the centre of the brain to the cortex) of 50 mm, but this distance will be slightly smaller in this paediatric cohort. Acquisitions were excluded if they had a mean framewise displacement >0.5 or if the maximum absolute displacement from the reference volume exceeded 4 mm. T1-weighted scans were visually assessed and those with severe movement artefact, which would affect the registration of the subject data to the standard template, were excluded.

### Group Independent Component Analysis

Following preprocessing, resting-state fMRI data for the whole cohort were analysed using spatial group independent component analysis (GICA). GICA decomposes data into maximally spatially independent components, whose time courses can be linearly combined to reconstruct the original data. GICA was applied using GIFT (Calhoun et al., 2001; Erhardt et al., 2011), as follows. An initial dimensionality reduction step was applied to the fMRI data for each subject, using principal component analysis (PCA) to reduce 295 timepoint data to 120 directions of maximal variability. Subject data for the whole cohort were then concatenated across time and a group PCA step reduced this into 100 components with the expectation maximisation algorithm. The infomax algorithm (Bell and Sejnowski, 1995) was then used to calculate 100 independent components from the reduced-dimensionality group data. To ensure robust estimation of independent components, ICA was repeated 20 times using ICASSO, and aggregate spatial maps were estimated as the modes of component clusters. We selected only components which gave a stability index (I_q_) >0.8 in ICASSO. For these components, subject-specific spatial maps and time courses were calculated using the GICA back-reconstruction method, which is analogous to dual regression, differing only in the projection through the initial PCA step (Erhardt et al., 2011).

We inspected the spatial maps and temporal properties of the independent components to identify intrinsic connectivity networks (ICNs) based on the criteria described by (Allen et al., 2014), as follows: a) peak activation coordinates were in grey matter and had low spatial overlap with known artifacts (vascular, ventricular, motion or susceptibility); b) time courses were dominated by low-frequency fluctuations, characterised by a high ratio of power <0.10 Hz to 0.15-0.25 Hz (Cordes et al., 2000); c) time courses had a high dynamic range (the difference between maximum and minimum power frequencies). Through this process we identified 31 ICNs which were sorted into seven functional networks (basal ganglia, sensorimotor, auditory, visual, DMN, attention/cognitive control, cerebellar) based on the spatial maps provided by (Shirer et al., 2012).

For these 31 ICNs, subject-specific time courses (obtained from back-reconstruction, as described above) were detrended (for linear, quadratic and cubic trends), despiked using AFNI’s 3dDespike algorithm (http://afni.nimh.nih.gov/afni) to replace outliers with values calculated from a third order spline fit to neighbouring clean data points, and low-pass filtered using a fifth order Butterworth filter with a 0.15 Hz cut-off frequency.

### Static Functional Connectivity

We calculated a 31 x 31 static functional connectivity (sFC) matrix for each subject by measuring the pairwise Pearson correlation coefficient between the subject-specific timeseries of each ICN and applying Fisher’s z-transform.

### Dynamic Functional Connectivity

Recent studies have demonstrated that investigating resting-state connectivity in shorter time windows of tens of seconds can reveal dynamic changes in FC, offering greater insight into functional properties of brain networks (Allen et al., 2014; Gonzalez-Castillo et al., 2015; Handwerker et al., 2012). We assessed time-varying dFC in this cohort using the methodology described in (Spencer and Goodfellow, 2022), which builds on the standard sliding-window correlations framework (Allen et al., 2014) by including a dimensionality reduction step prior to clustering. We used deep clustering (Caron et al., 2018; Guo et al., 2017), which consists of autoencoders for dimensionality reduction prior to k-means clustering, as this provides more accurate measurements of state temporal properties in synthetic data than other dimensionality reduction methods, or k-means clustering alone (Spencer and Goodfellow, 2022). Autoencoders are a type of artificial neural network which, in dimensionality reduction applications, are trained to copy the input data to the output via a low-dimensional encoding layer (Goodfellow et al., 2016; Vincent et al., 2008). The low-dimensional encoding layer extracts salient features from which the original data can be reproduced via the decoding layers (Guo et al., 2017; Xie et al., 2016). Sliding-window correlations and deep clustering were performed as follows.

First, we used the sliding-window correlations approach to convert each ICN time course for each subject to a series of FC matrices, representing time-varying functional connections (Figure 1). We used a tapered window of length 50 TR (45.3 s), created by convolving a rectangular window with a Gaussian function with a sigma of 6 TR (5.436 s). Sliding the window in steps of 1 TR (0.906 s), we calculated FC within each window by estimating covariance from the precision matrix with L1 regularisation (Allen et al., 2014; Smith et al., 2011; Varoquaux et al., 2010), where the regularisation parameter, λ_L1_, was estimated for each subject using cross-validation, and applied Fisher’s z-transform.

**Figure 1:**
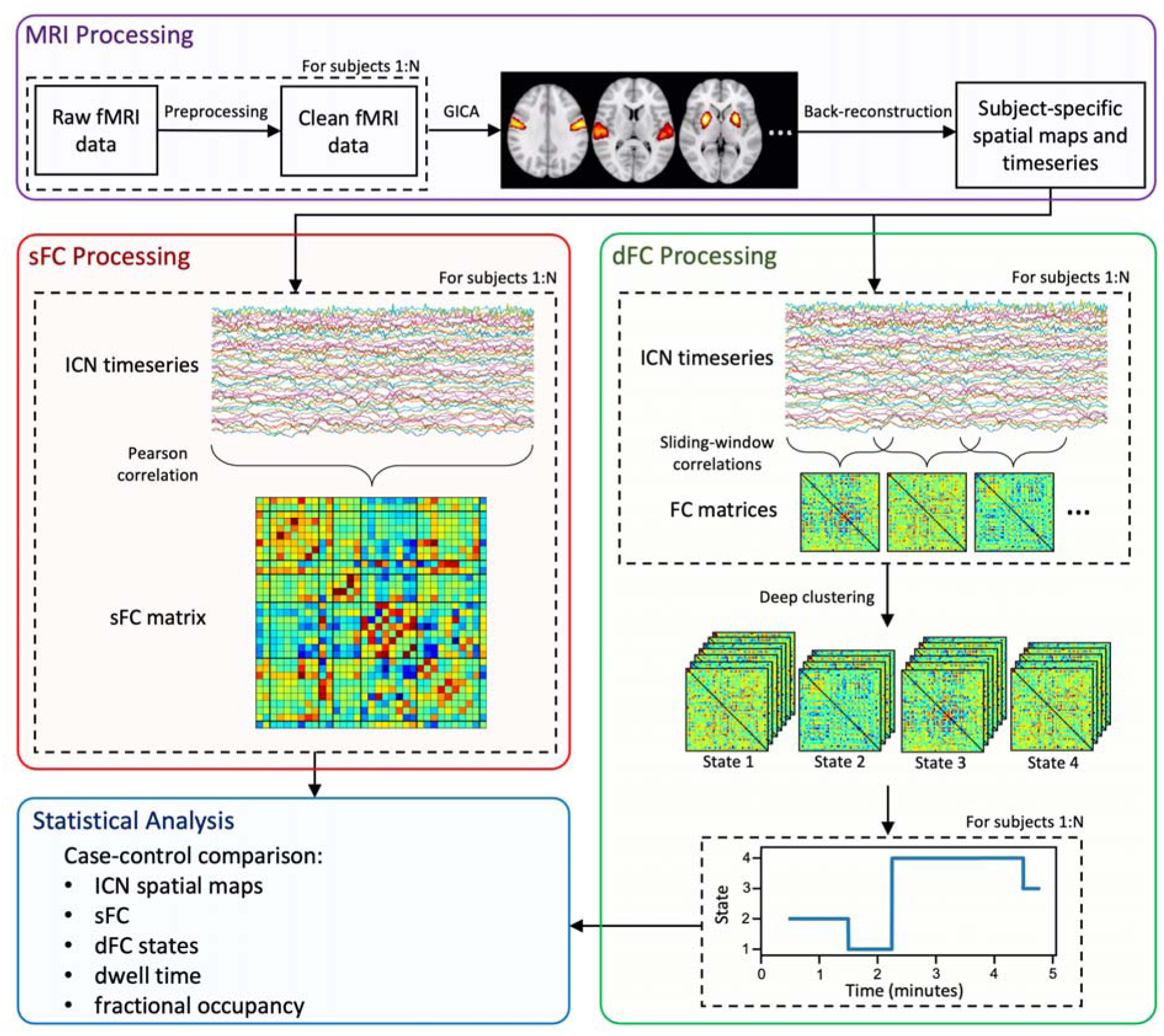
Pipeline of analysis methods. Each subject’s resting-state fMRI data was preprocessed, then group independent component analysis (GICA) was used to extract intrinsic connectivity networks (ICNs). We identified 31 ICNs and obtained subject-specific spatial maps and time courses using back-reconstruction. Static functional connectivity (sFC) was computed for each subject by measuring pairwise correlation between ICNs. Dynamic functional connectivity (dFC) was computed by sliding-window correlations followed by deep clustering (Spencer and Goodfellow, 2022) to group FC windows into k = 4 states (determined using the elbow criterion of the within-cluster distance to the between-cluster distance). Dwell time and fractional occupancy were measured for each subject. ICN spatial maps and characteristics of sFC and dFC were compared between cases and controls.

For dimensionality reduction, we used the autoencoder architecture described in (Spencer and Goodfellow, 2022). Specifically, this consisted of a fully-connected autoencoder with three encoding layers (number of units: 512, 256, 32) and a symmetric decoder. Linear activation functions were used for the low-dimensional layer and output layer, and rectified linear unit (ReLU) activation functions were used for all other layers. We trained the autoencoder for 200 epochs with a batch size of 50, using the Adam optimiser (Kingma and Ba, 2014) to minimise the mean-squared error (MSE) between the input and output.

We then applied k-means clustering to the low-dimensional representation of the dFC data for all subjects, as follows. First we selected exemplar FC windows at local maxima in variance and applied 128 repetitions of k-means (max 1000 iterations) to the low-dimensional representation of these windows, each initialised with the k-means++ algorithm (Arthur and Vassilvitskii, 2006). From these 128 runs, the set of centroids which gave the lowest sum of squared error between each data point and its nearest centroid was used to initialise k-means clustering (max 10,000 iterations) for all windows.

We determined the number of clusters using the elbow criterion of the within-cluster distance to the between-cluster distance, which resulted in k = 4. For each subject, we measured the mean dwell time of each cluster (the average time spent in that state) and the fractional occupancy of each cluster (the fraction of the total scan time spent in that state).

### Statistical Analysis

After data processing, for each subject we had: i) subject-specific spatial maps for 31 ICNs; ii) a subject-specific sFC matrix denoting pairwise FC between ICNs; and iii) dFC outputs for each state, consisting of state FC matrices and measurements of dwell time and fractional occupancy.

To investigate differences in ICN spatial maps between cases and controls, we performed case-control comparison of subject-specific spatial maps for each ICN using FSL’s RANDOMISE (Winkler et al., 2014). Age and sex were included as covariates in a general linear model, performing two-tailed voxelwise comparison between cases and controls with 10,000 permutations and applying threshold-free cluster enhancement to control the family-wise error rate.

We then investigated group differences in sFC between cases and controls; we regressed age and sex from each pairwise functional connection (pairwise association between ICNs), and performed a two-tailed t-test using the residuals. We present uncorrected results, in addition to results after applying FDR correction for multiple comparisons.

To compare dFC characteristics, we compared dwell time and fractional occupancy between cases and controls using ANCOVA with age and sex included as covariates. To assess group differences in state FC matrices between cases and controls, we calculated subject-specific state FC matrices as the median of FC windows assigned to each state for a given subject. We performed element-wise comparison between cases and controls by first regressing age and sex from each functional connection, then performing a two-tailed t-test using the residuals. We applied FDR multiple comparisons correction.

## Results

### Participant Demographics

50 cases and 43 controls were recruited for the CoolMRI study. 7 cases and 4 controls did not want to undergo scanning and 7 cases had incomplete data due to movement during the scan. Quality control of the fMRI data resulted in rejection of 13 cases and 19 controls. One additional case was rejected due to poor quality of their T1-weighted image, meaning that the data could not be spatially normalised. This left 22 cases and 20 controls with suitable data. Participant demographics are shown in Table 1. There was no significant difference between cases and controls in age, sex, deprivation index, or framewise displacement. As previously reported (Spencer et al., 2021a, 2021b), cases had lower cognitive scores (p = 0.0053) measured by the Wechsler Intelligence Scale for Children 4th Edition (Kaufman et al., 2006), and a larger proportion of the case group were at risk of motor impairment (p = 0.0221), defined as a score under the 15^th^ centile on the Movement Assessment Battery for Children 2nd Edition (MABC-2) (Henderson et al., 2007).

**Table 1:**
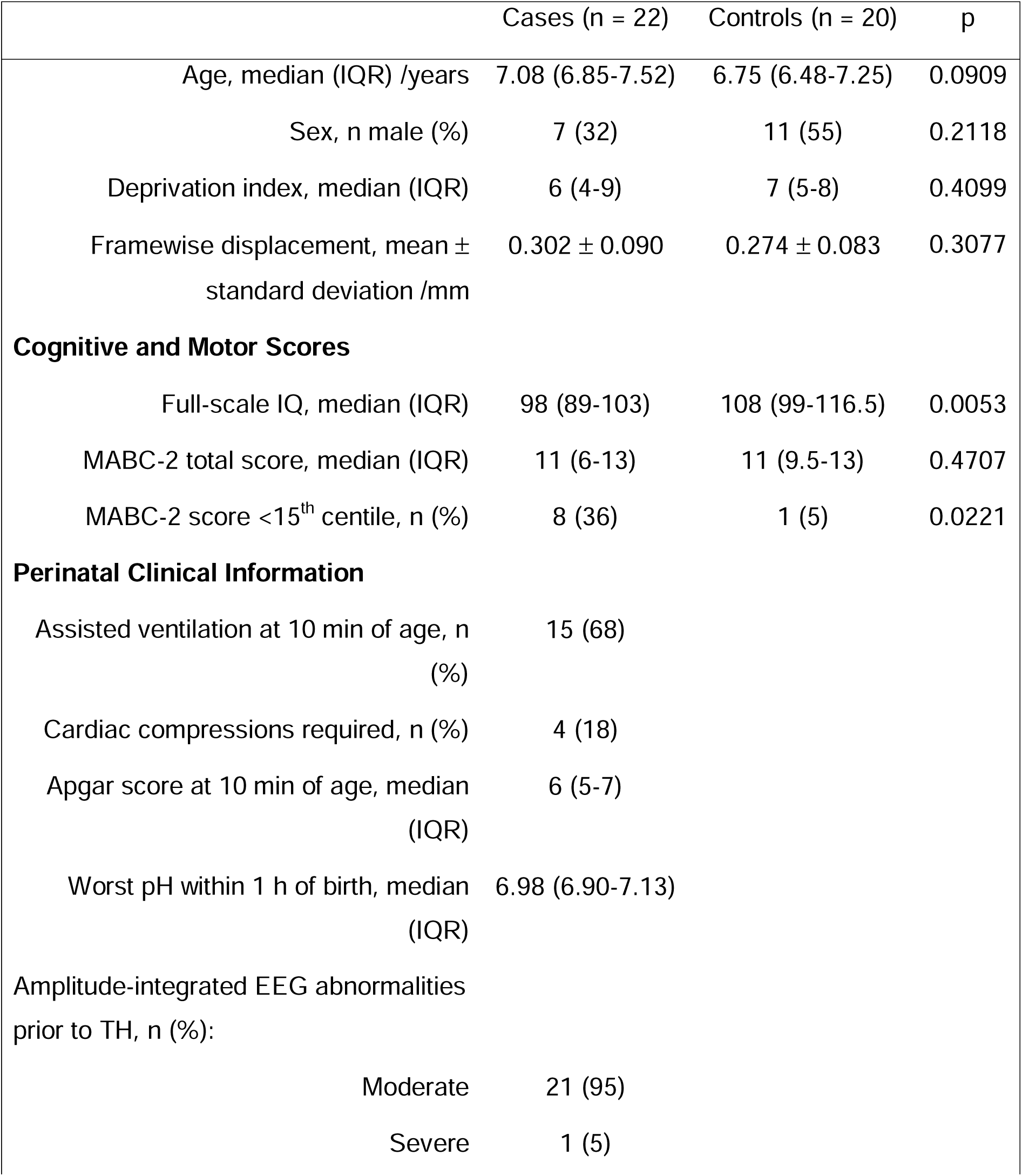
Participant demographics and perinatal clinical information. Apgar score is measured on a 1-10 scale where a higher score indicates healthier (7-10 indicates good health). Perinatal asphyxia is characterised by pH <7.20.

|  | Cases (n = 22) | Controls (n = 20) | p |
| --- | --- | --- | --- |
| Age, median (IQR) /years | 7.08 (6.85-7.52) | 6.75 (6.48-7.25) | 0.0909 |
| Sex, n male (%) | 7 (32) | 11 (55) | 0.2118 |
| Deprivation index, median (IQR) | 6 (4-9) | 7 (5-8) | 0.4099 |
| Framewise displacement, mean ± standard deviation /mm | 0.302 ± 0.090 | 0.274 ± 0.083 | 0.3077 |
| **Cognitive and Motor Scores** |  |  |  |
| Full-scale IQ, median (IQR) | 98 (89-103) | 108 (99-116.5) | 0.0053 |
| MABC-2 total score, median (IQR) | 11 (6-13) | 11 (9.5-13) | 0.4707 |
| MABC-2 score <15<sup>th</sup> centile, n (%) | 8 (36) | 1 (5) | 0.0221 |
| **Perinatal Clinical Information** |  |  |  |
| Assisted ventilation at 10 min of age, n (%) | 15 (68) |  |  |
| Cardiac compressions required, n (%) | 4 (18) |  |  |
| Apgar score at 10 min of age, median (IQR) | 6 (5-7) |  |  |
| Worst pH within 1 h of birth, median (IQR) | 6.98 (6.90-7.13) |  |  |
| Amplitude-integrated EEG abnormalities prior to TH, n (%): |  |  |  |
| Moderate | 21 (95) |  |  |
| Severe | 1 (5) |  |  |

### Intrinsic Connectivity Networks

Figure 2 shows the spatial maps of the 31 ICNs identified from independent component analysis, grouped into seven functional networks (Shirer et al., 2012). These ICNs are consistent with those found in previous studies of children (de Bie et al., 2012; Muetzel et al., 2016; Rashid et al., 2018; Thomason et al., 2011) and adults (Allen et al., 2014; Damaraju et al., 2014; Fiorenzato et al., 2019; Shirer et al., 2012). Details of each independent component are provided in Supplementary Table 1, with spatial maps shown in Supplementary Figure 1. There were no case-control differences in ICN spatial maps (p>0.05) indicating the spatial extent of independent components are consistent between groups.

**Figure 2:**
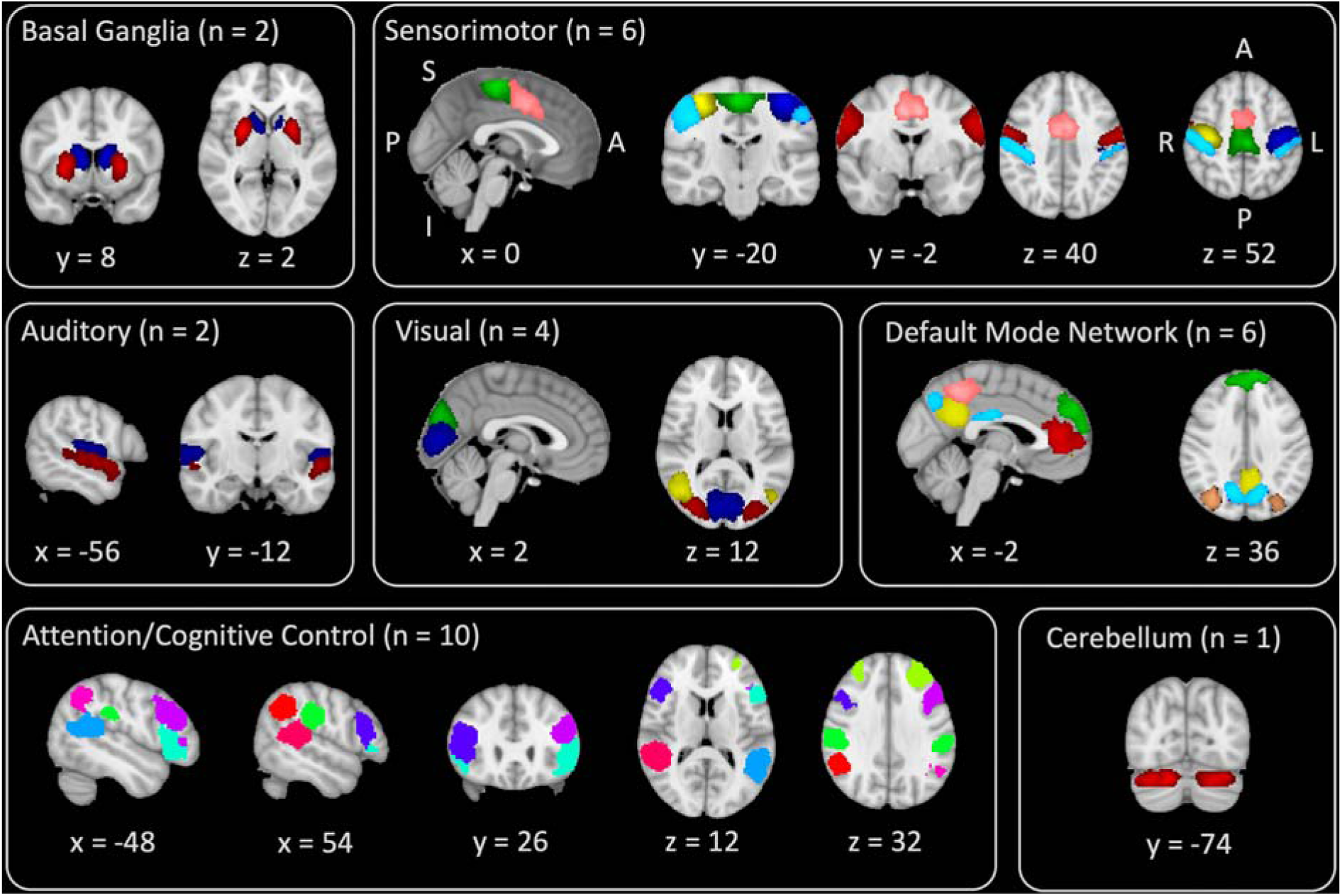
Spatial maps of intrinsic connectivity networks identified by group independent component analysis, grouped into functional networks, with arbitrary colours for visualisation. Orientation is indicated by the labels in the sensorimotor panel as follows: S/I = superior/inferior; A/P = anterior/posterior; R/L = right/left.

### Static Functional Connectivity

The average sFC matrix for the whole cohort is shown in Figure 3. Similar to previous studies (Allen et al., 2014; Rashid et al., 2018), sFC patterns in this cohort show modular organisation, with most functional networks (e.g. sensorimotor, visual, DMN, attention/cognitive control) exhibiting positive connectivity between ICNs within the network. The ICNs which comprise the DMN exhibited negative correlation with most other functional networks.

**Figure 3:**
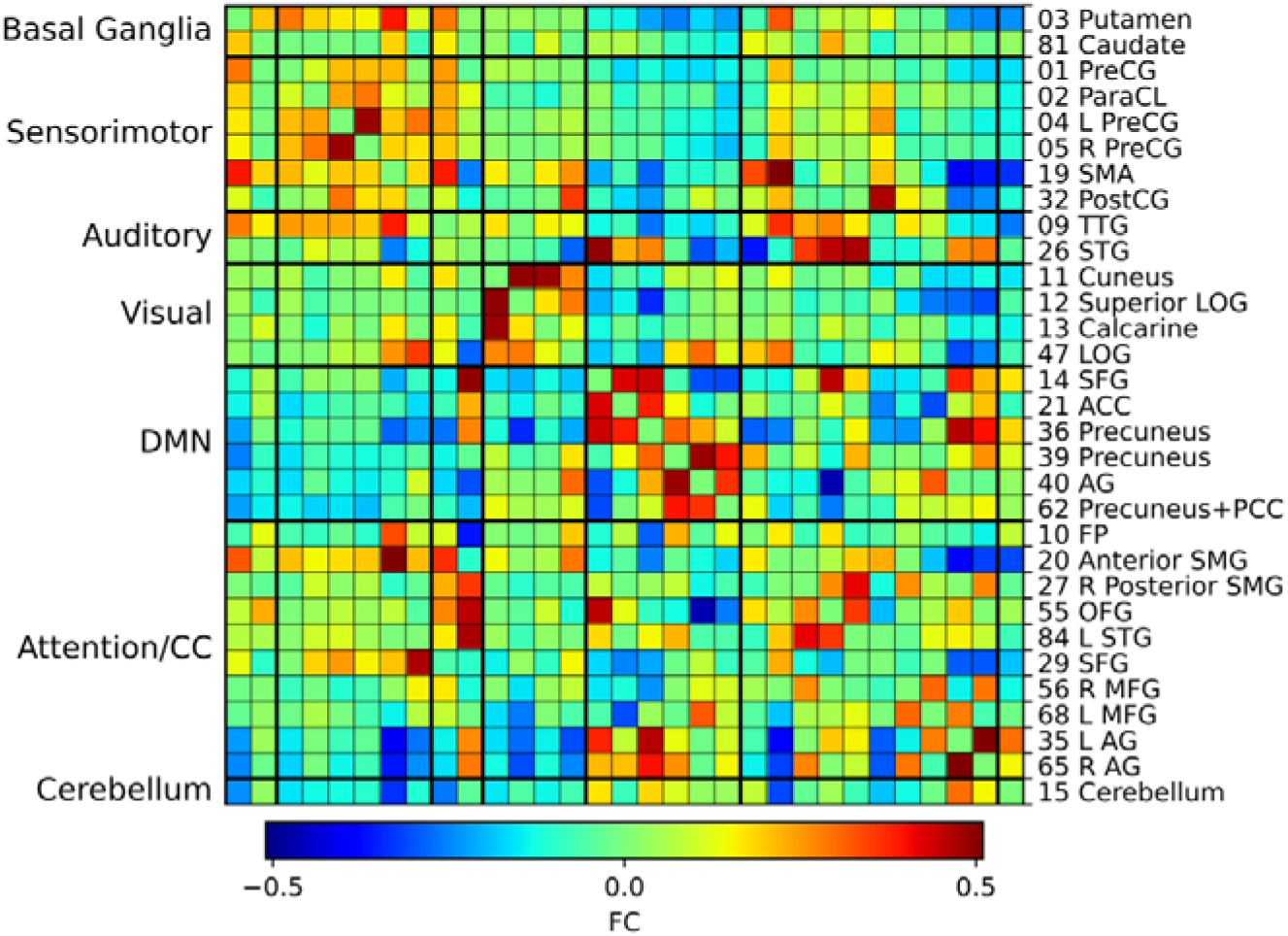
Average static functional connectivity (sFC) matrix for the whole cohort. Independent component number and label is shown in the right, corresponding to the intrinsic connectivity networks (ICNs) shown in Supplementary Table 1. ICNs are arranged into seven functional networks shown on the left. Pairwise functional connectivity (FC) is indicated by the colour bar. The number of each component corresponds to the independent component number in Supplementary Table 1 and Supplementary Figure 1.

We investigated group differences in sFC between ICNs after regressing age and sex. After FDR correction there were no case-control differences in sFC. The uncorrected t-statistic map is presented in Figure 4. Before multiple comparisons correction, there were group differences in FC within the attention/cognitive control network, and between this and other functional networks (Figure 4).

**Figure 4:**
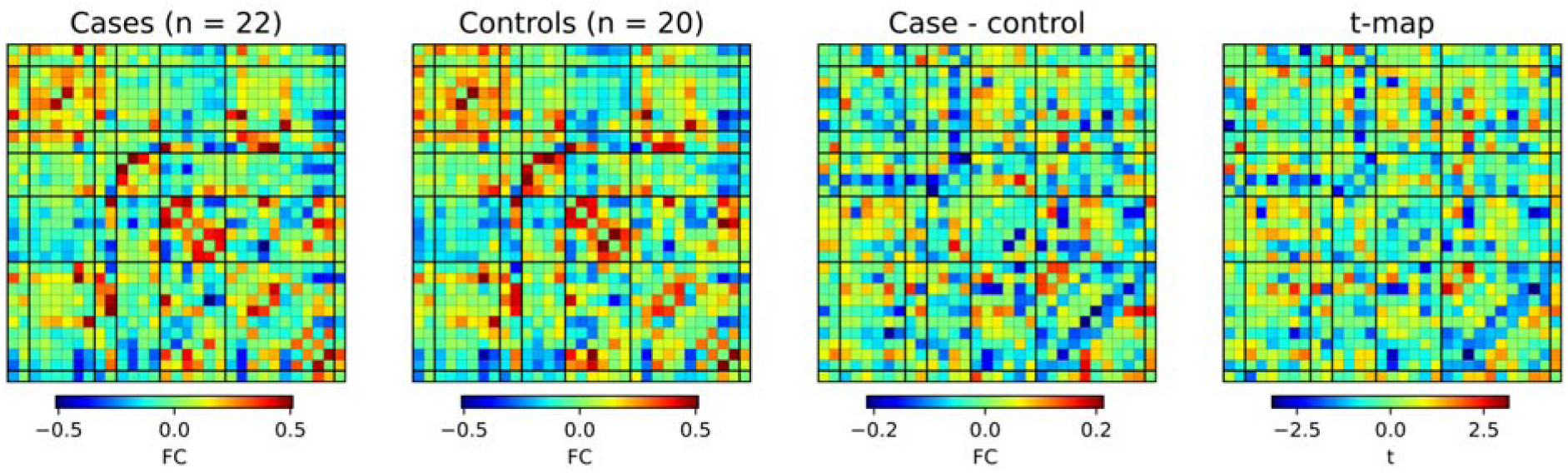
Case-control differences in static functional connectivity (sFC). Colourmaps from left to right show average sFC in cases, average sFC in controls, the difference between these, and the t-statistic from a two-tailed t-test of residual functional connectivity (FC) after regressing age and sex. A t-statistic of |t| > 2.02 corresponds to uncorrected p < 0.05. None of these differences were significant after FDR-correction.

### Dynamic Functional Connectivity

We used sliding-window correlations and deep clustering to identify k = 4 repetitively occurring FC states, shown in Figure 5 along with the distribution of residual dwell time and fractional occupancy of each state in cases and controls after regressing age and sex. State 1, which makes up the largest proportion of FC windows, is characterised by very weak connectivity among most ICNs. Previous studies have found similar connectivity patterns in the most frequently observed state, and have suggested this may be the average of multiple additional states which are not sufficiently distinct or prevalent to be distinguished (Allen et al., 2014; Marusak et al., 2017). State 2 is characterised by positive connectivity within the sensorimotor network, within the DMN, and between the sensorimotor and attention/cognitive control networks, but negative connectivity between the DMN and other functional networks. State 3 exhibits strong positive connectivity between ICNs in the visual network, and between ICNs in the DMN, but strong negative connections between many ICNs across all networks. State 4 represents a highly integrated state, characterised by positive connectivity between ICNs across all networks. After FDR-correction there were no differences in the state FC matrices. There were no case-control differences in dwell time or fractional occupancy in any of the states.

**Figure 5:**
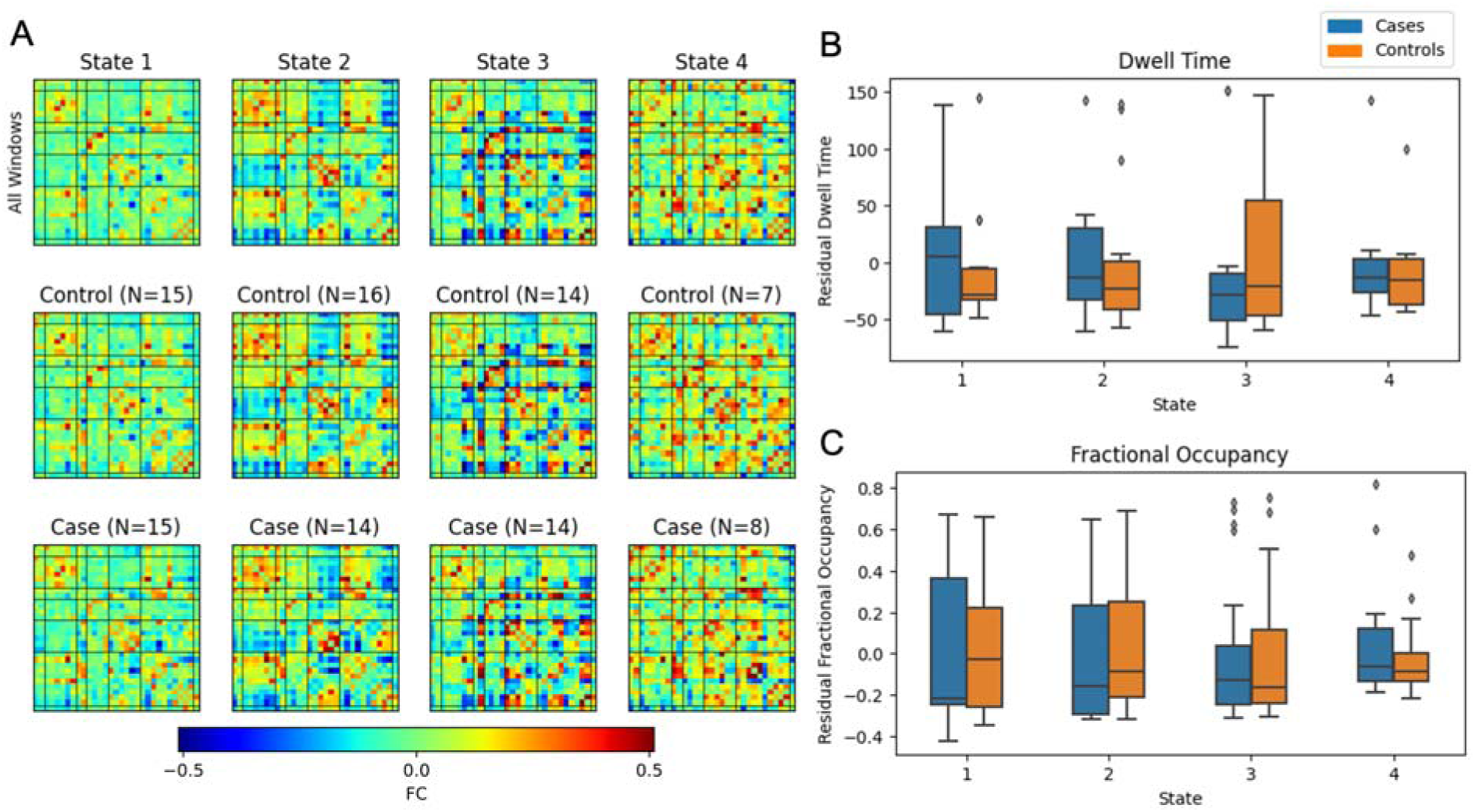
Dynamic functional connectivity (dFC) state maps and temporal properties. A) dFC state maps, ordered by prevalence, are shown for the whole cohort (top row), and for controls (middle) and cases (bottom). The distribution of residual dwell time (B) and fractional occupancy (C), after regressing age and sex, are shown as box plots with boxes indicating the interquartile range with a line for the median and whiskers extending to the range of the data, not including outliers which are shown as diamonds.

## Discussion

In this study we investigated resting-state networks measured from fMRI in children treated with therapeutic hypothermia for HIE, who did not develop cerebral palsy, and controls matched for age, sex and socioeconomic status. There were no case-control differences in ICN spatial maps, sFC between ICNs, and dFC states and temporal characteristics. From 100 independent components derived by spatial group ICA, we identified 31 ICNs based on characteristics of the time courses, spatial maps and power spectra. These ICNs correspond to known resting-state networks previously reported in both children and adults (Allen et al., 2014; de Bie et al., 2012; Marusak et al., 2017; Muetzel et al., 2016; Rashid et al., 2018; Shirer et al., 2012; Thomason et al., 2011). We found no case-control differences in the spatial maps of these ICNs. We investigated sFC by measuring pairwise correlations between ICN time courses over the duration of the scan for each subject. Before multiple comparisons correction there were case-control differences in attention and cognitive networks, however these were not significant after FDR correction. Using dFC analysis to investigate dynamic fluctuations in resting-state activity revealed k = 4 repetitively occurring brain states. There were no differences between cases and controls in dwell time, fractional occupancy, or state FC matrices.

There have been few studies of resting-state FC in children with HIE (Smyser et al., 2019); resting-state networks in children with HIE have previously only been examined in the neonatal period. Jiang et al. investigated resting-state FC in motor networks in neonates cooled for HIE (5 mild, 8 moderate and 3 severe, as determined using Sarnat criteria), in comparison with healthy controls at 1-2 weeks of age (Jiang et al., 2022). They reported reduced FC between primary motor regions in neonates with HIE, and case-control differences in FC spatial maps. Tusor et al. reported reduced FC in auditory, somatomotor, visual and default-mode networks in infants cooled for HIE compared to healthy controls (Tusor, 2014). In a retrospective study of neonates with acute brain injury, 27 of whom were cooled for HIE (14 mild, 7 moderate, 6 severe as determined by Sarnat criteria), more severe outcomes were associated with atypical resting-state activity in the basal ganglia, frontoparietal and default-mode networks (Boerwinkle et al., 2022). Additionally, Li et al. found that functional brain networks in neonates with severe HIE had lower local efficiency and clustering coefficient compared to those with moderate HIE at around 2 weeks of age, indicating reduced capacity for segregated functional processing (Li et al., 2019). However, the authors did not report whether participants received therapeutic hypothermia and it is not standard care nationwide in China, where the study was carried out (Wang et al., 2021). Our cohort did not include those with cerebral palsy, thus is not directly comparable to the previous studies on infants too young to rule out a diagnosis of cerebral palsy. Our cohort was almost entirely made up of those with moderate HIE (only one case had severe HIE); it is possible that a cohort made up of cases with severe HIE might have more distinguishable differences in FC. However, in the same cohort with a similar proportion of severe vs moderate HIE, we previously reported widespread alterations to structural connectivity and white matter diffusion properties (Spencer et al., 2021b).

The limited case-control differences in ICN spatial maps, sFC, and dFC characteristics between the case groups and matched healthy controls is despite previous findings in the same cohort showing widespread alterations to brain structure and white matter connectivity, which are associated with cognitive and motor impairments in cases (Byrne et al., 2023; Robb et al., 2022; Spencer et al., 2023, 2022, 2021b, 2021a). This may be due to the small sample size; our previous work has identified heterogeneity in the severity of impairments to brain structure and cognition in this cohort (Lee-Kelland et al., 2020; Spencer et al., 2022, 2021b), therefore any alterations to resting-state brain activity are also likely to be heterogeneous. The subtle differences shared across the cohort would require a large sample size to distinguish from healthy resting-state activity. Before multiple comparisons correction, there were group differences in FC between the attention/cognitive control network and the sensorimotor and visual networks (Supplementary Figure 2). This may reflect neural correlates of the attention and visuospatial processing difficulties observed in behavioural studies in this cohort (Tonks et al., 2019), and the altered structural connectivity to regions association with attention and visuospatial processing previously reported (Spencer et al., 2021b). However, further study with a larger sample size is required to robustly identify these differences. Differences in brain activity in this cohort may also be detected by a task-based fMRI paradigm which demands the specific aspects of cognition known to differ between cooled children and controls (Zhao et al., 2023).

Given the difficulty in recruiting and studying patients who were cooled for HIE up to eight years ago, such exploratory studies of whole-brain functional connectivity are always likely to be underpowered to detect results corrected for family wise error. As such we took a pragmatic approach of presenting the data with and without correction, pointing out overlap with previous findings (which were corrected for multiple comparisons), and suggest that by correcting for the sheer number of ICNs observed makes it difficult to meet the most stringent statistical tests.

It is possible that the minimal group differences reflect a level of recovery of healthy resting-state brain function, despite the structural differences in this cohort. This may suggest that healthy cognitive function could also be recovered in this developmental period. For example, if the appropriate support or intervention was provided in this developmental period between infancy and early school-age, it may be possible to minimise cognitive impairments (Astle et al., 2015; Blasco et al., 2023; Galetto and Sacco, 2017; Prosperini et al., 2015).

### Strengths & Limitations

To our knowledge, this is the first study to investigate resting-state FC in children with HIE beyond the neonatal period, and the first to investigate dFC in any children with HIE. The main difficulty when scanning children of this age group is movement during the scan, which can affect FC measurements (Lurie et al., 2020; Power et al., 2015; Van Dijk et al., 2012). We took steps to alleviate the effect of movement, using thorough preprocessing and data cleaning procedures to identify and regress noisy signals and motion parameters, in addition to rejecting participants based on quantitative evaluation of movement during the scan. As a result, there was no group difference in framewise displacement. However, rejection of those with excessive movement resulted in a small sample size, which is the main limitation of this study.

## Data Availability

The data that support the findings of this study are available from the corresponding author, upon reasonable request. The code used for dFC analysis (including sliding-window correlations and deep clustering) is available at GitHub (https://github.com/apcspencer/dFC_DimReduction) (Spencer and Goodfellow, 2022).

## Acknowledgements

We would like to thank the children and their families for participating, Ngoc Jade Thai for her assistance with MR sequences, Aileen Wilson for her radiographical expertise, and Marianne Thoresen, Frances M Cowan, Hollie Byrne and Richard Lee-Kelland for their role in the CoolMRI study.

