## Supplementary Materials for "Resting-state functional connectivity in children cooled for neonatal encephalopathy"

| | | | | Peak coordinates | | |
|---|---|---|---|---|---|---|
| IC | Region | $Z_{max}$ | N voxels | x (mm) | y (mm) | z (mm) |
| Basal Ganglia (n = 2) | | | | | | |
| IC03 | R putamen | 12.3 | 1070 | 28 | 2 | 0 |
| | L putamen | 10.7 | 919 | -24 | 6 | 2 |
| IC81 | Bi caudate | 9.36 | 1866 | 12 | 12 | 14 |
| Sensorimotor (n = 6) | | | | | | |
| IC01 | L precentral gyrus | 9.84 | 1158 | -56 | -4 | 32 |
| | R precentral gyrus | 9.21 | 1127 | 50 | -6 | 32 |
| IC02 | Bi paracentral lobule | 10.4 | 2025 | 4 | -22 | 58 |
| IC04 | L precentral gyrus | 11.0 | 1635 | 11 | -40 | 56 |
| | R cerebellum | 6.06 | 124 | 22 | -48 | -20 |
| IC05 | R precentral gyrus | 12.1 | 1383 | 36 | -20 | 56 |
| IC19 | Bi supplementary motor area | 8.44 | 1740 | 8 | -4 | 44 |
| IC32 | R postcentral gyrus | 8.73 | 1512 | 42 | -26 | 46 |
| | L postcentral gyrus | 6.04 | 599 | -40 | -34 | 54 |
| Auditory (n = 2) | | | | | | |
| IC09 | R transverse temporal gyrus | 8.21 | 1318 | 52 | -18 | 8 |
| | L transverse temporal gyrus | 7.42 | 1024 | -50 | -24 | 12 |
| IC26 | L superior temporal gyrus | 7.41 | 1376 | -54 | -12 | -4 |
| | R superior temporal gyrus | 5.15 | 250 | 58 | 2 | -10 |
| Visual (n = 4) | | | | | | |
| IC11 | Bi cuneus | 8.86 | 2205 | 4 | -82 | 28 |
| IC12 | R lateral occipital gyrus | 8.25 | 1297 | 30 | -80 | 14 |
| | L lateral occipital gyrus | 7.16 | 926 | -30 | -84 | 14 |
| IC13 | Bi calcarine | 9.07 | 2450 | -2 | -82 | 4 |
| IC47 | R lateral occipital gyrus | 9.03 | 1290 | 46 | -64 | 10 |
| | L lateral occipital gyrus | 5.62 | 148 | -46 | -72 | 12 |
| DMN (n = 6) | | | | | | |
| IC14 | Bi superior frontal gyrus | 6.87 | 1836 | -8 | 56 | 36 |
| IC21 | Bi anterior cingulate | 8.10 | 2198 | -2 | 40 | 12 |
| IC36 | Bi precuneus | 7.64 | 1428 | -4 | -48 | 28 |
| IC39 | Bi precuneus | 7.55 | 1623 | -2 | -50 | 50 |
| IC40 | R angular gyrus | 6.59 | 756 | 34 | -68 | 36 |
| | L angular gyrus | 6.16 | 542 | -30 | -70 | 30 |
| IC62 | Bi precuneus | 8.21 | 1585 | 14 | -66 | 38 |
| | Bi posterior cingulate | 6.96 | 457 | -4 | -18 | 28 |
| Attention/cognitive control (n = 10) | | | | | | |
| IC10 | L frontal pole | 8.15 | 1484 | -28 | 44 | 34 |
| | R frontal pole | 5.81 | 382 | 32 | 46 | 36 |
| IC20 | R anterior supramarginal gyrus | 7.83 | 1074 | 54 | -26 | 28 |
| | L anterior supramarginal gyrus | 6.74 | 551 | -54 | -26 | 26 |
| IC27 | R posterior supramarginal gyrus | 9.54 | 1554 | 50 | -40 | 12 |
| IC55 | L orbitofrontal gyrus | 7.39 | 1171 | -46 | 28 | -4 |
| | R orbitofrontal gyrus | 5.9 | 346 | 48 | 32 | -4 |
| IC84 | L superior temporal gyrus | 7.55 | 1600 | -54 | -46 | 16 |
| IC29 | L superior frontal gyrus | 8.58 | 825 | -18 | -8 | 56 |
| | R superior frontal gyrus | 7.62 | 606 | 16 | -8 | 56 |
| IC56 | R middle frontal gyrus | 6.89 | 1952 | 46 | 22 | 18 |
| IC68 | L middle frontal gyrus | 7.54 | 1442 | -40 | 26 | 20 |
| IC35 | L angular gyrus | 5.66 | 654 | -48 | -56 | 42 |

| | | | | | | |
|---|---|---|---|---|---|---|
| | L posterior middle temporal gyrus | 5.92 | 363 | -62 | -24 | -12 |
| | L middle frontal gyrus | 5.68 | 234 | -38 | 18 | 46 |
| IC65 | R angular gyrus | 6.50 | 1022 | 52 | -50 | 38 |
| | R posterior middle temporal gyrus | 5.28 | 328 | 64 | -30 | -2 |
| Cerebellum (n = 1) | | | | | | |
| IC14 | Bi cerebellum | 8.11 | 1836 | 18 | -76 | -30 |

Supplementary Table 1: Coordinates of intrinsic connectivity networks (ICNs) identified by group independent component analysis. We identified 33 ICNs from the 100 ICs, which are grouped into seven functional networks. Spatial maps were converted to z-score and thresholded at z > 4 (displayed in Supplementary Figure 1). Maximum z-score, and peak coordinates (MNI standard space) are shown for clusters greater than 100 voxels.

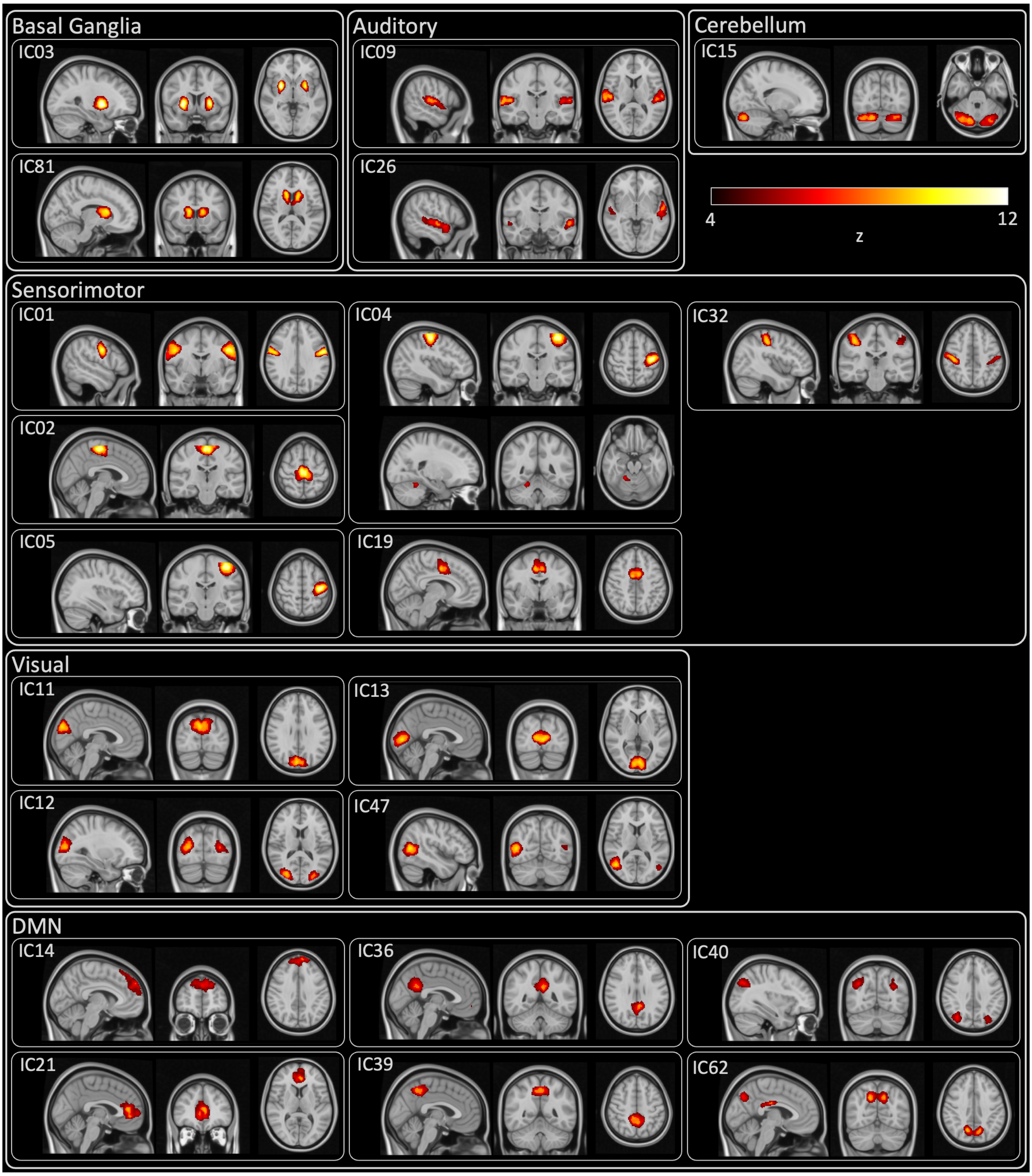


Supplementary Figure 1 (continues on next page).

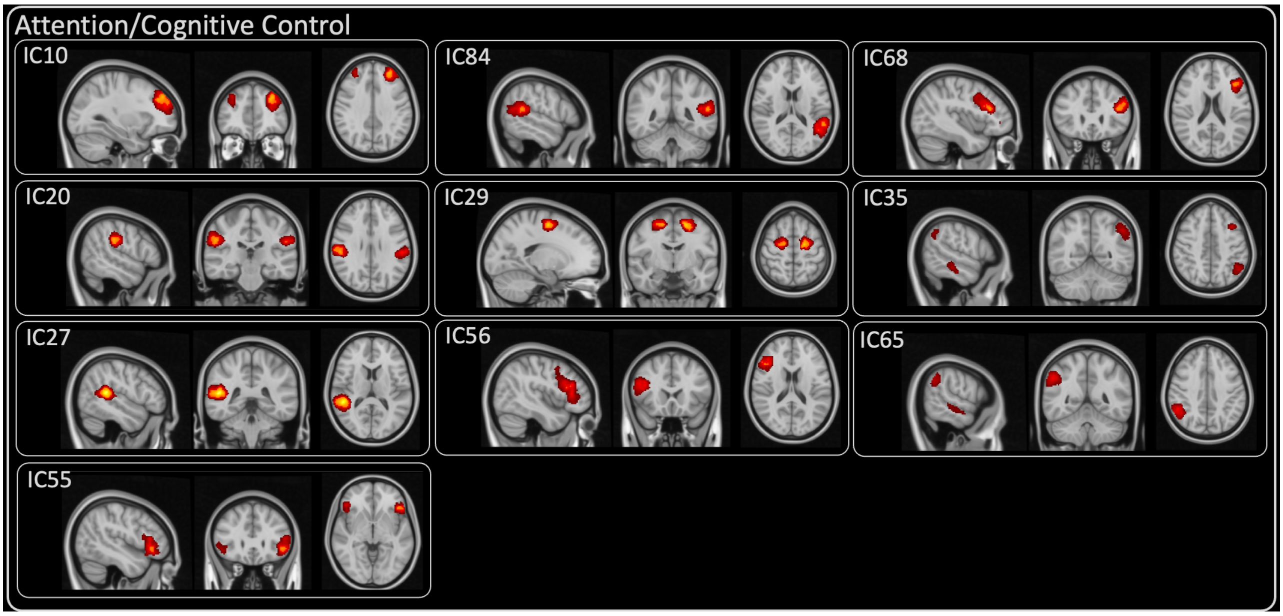

Supplementary Figure 1 (continued): Spatial maps of each intrinsic connectivity network (ICN). Spatial maps were converted to z-score and thresholded at z > 4, and are displayed on the MNI standard template. Coordinates, peak z-score, and cluster sizes are shown in Supplementary Table 1. Z-score is indicated by the colour bar.
